# Understanding Barriers to Clinical Trial Participation Among U.S. Women: A National Survey Study

**DOI:** 10.1101/2025.10.28.25337741

**Authors:** Doruntina Fida, Therese A. Rajasekera, Ancella Roy, Julia C. Wilson, Aleta Wiley, Ruth Lederman, Primavera A. Spagnolo

**Affiliations:** Connors Center for Women’s Health Research, Brigham and Women’s Hospital, Boston, MA, USA; Department of Psychiatry, Brigham and Women’s Hospital, Boston, MA, USA; Harvard Medical School, Boston, MA, USA; University of Massachusetts Chan Medical School, Worcester, MA, USA; Columbia University Vagelos College of Physicians and Surgeons, New York, NY 10032; Survey and Data Management Core, Dana Farber Cancer Institute, Boston, MA

**Author notes:** Correspondence should be addressed to: Primavera A Spagnolo, MD, PhD, Mary Horrigan Connors Center for Women’s Health Research Dept. of Psychiatry, Brigham and Women’s Hospital, Harvard Medical School, Thorn Building, 75 Francis Street, Boston, MA.

**Keywords:** Clinical trial participation, Women’s Health, Health disparities, Race and ethnicity, Sex differences

## Abstract

Despite the persistent underrepresentation of women—particularly those from racially and ethnically minoritized groups—in clinical research, little is known about their perspectives on participation. This study examined healthcare experiences, access, and attitudes toward clinical trials among U.S. women and assessed how race, socioeconomic status, and healthcare access intersect to shape willingness to participate (WTP). We conducted a national cross-sectional online survey (January–March 2023) of 5,301 women aged 18–70 years. The 81-item questionnaire assessed demographics, health status, healthcare access, and clinical trial experiences. Among 4,987 respondents reporting race (77% White, 14% Black, 7% Asian, 2% Other), nearly 80% expressed interest in participating in clinical trials, yet only 11% had been invited and 7% had enrolled. In adjusted models, WTP was lower among Black (β = −0.06; P = .04) and Asian (β = −0.09; P = .01) women than among White women, whereas higher educational attainment and multimorbidity predicted greater WTP. Altruism, clear study explanations, and financial compensation were key motivators, while time burden and concerns about side effects were major barriers, with the salience of these factors varying by race. Most respondents (88%) endorsed the importance of women’s inclusion and sex-specific reporting, though neutrality on these issues was more frequent among racially minoritized women. Despite high interest, structural and informational barriers continue to constrain women’s engagement in clinical research, underscoring the need for trust-building, burden-reducing, and culturally responsive strategies to promote equitable participation and improve representation across racial groups.

## Introduction

For decades, clinical guidelines, diagnostics, and therapeutics have been developed primarily based on data from male populations, with limited attention to biological sex differences or the unique healthcare needs of women^1^. These systemic oversights have resulted in significant knowledge gaps in the prevention, diagnosis, and treatment of conditions that affect women differently or disproportionately^2^. Moreover, diseases that exclusively affect women − such as endometriosis and gynecologic cancers − have been largely understudied^3^. As a result, many women continue to receive care that is less precise, less effective, and often delayed, leading to poorer outcomes across a wide range of health conditions^4^.

A major contributor to these disparities is the persistent underrepresentation of women in clinical research, despite multinational policy and regulatory initiatives over the past decades designed to promote inclusion^5–10^. Numerous studies have consistently demonstrated that women’s enrollment in clinical trials lags behind both parity (equal representation of both sexes) and proportionality (representation relative to disease prevalence), across multiple therapeutic areas^11–15^. Disparities are further amplified among racially and ethnically minoritized women, who remain disproportionately excluded from FDA-regulated trials, post-marketing studies, comparative effectiveness trials, and vaccine trials [for a review see ^16^]. Older women are also frequently under-enrolled, limiting evidence regarding treatment safety and efficacy in this population^17^. Furthermore, even when women are included, data are rarely disaggregated or analyzed by sex, precluding rigorous evaluation of sex-specific effects^18,19^.

In response to these gaps, an expanding literature has begun to examine determinants of research participation among women, with increased attention to individuals’ perspectives and experiences. A systematic review of 63 studies comparing willingness to participate (WTP) in clinical trials by sex found lower WTP among women than men, driven by three recurrent influences: (i) the social and caregiving burden of participation; (ii) heightened risk perception and mistrust; and (iii) the quality of interactions with the research team^20^. Conversely, practical, low-burden strategies—such as transportation/parking support and telephone follow-up—were associated with higher WTP among women. Complementing this work, women-focused investigations have evaluated additional determinants, particularly race/ethnicity and reproductive stage, to delineate how these factors shape barriers to participation and trial experiences. Despite valuable insights, this literature often relies on disease-specific cohorts or samples limited to discrete reproductive stages, constraining generalizability^21–24^. Moreover, racial/ethnic influences are frequently examined within a single group^25–30^, hindering assessment of how racial context intersects with sex to shape participation. Finally, there is a dearth of studies assessing women’s awareness that biological sex influences the safety and efficacy of therapeutics, and why women’s inclusion in clinical trials is therefore essential.

To address these gaps, we conducted a national survey of a racially diverse sample of U.S. women to assess interest in, attitudes toward, and experiences with clinical trials and to test awareness of sex-based differences in relation to research engagement. Because participation is shaped by social and structural determinants, we examined racial differences alongside related factors—socioeconomic status, access to healthcare, and self-reported health—to delineate how these intersect to contribute to disparities in women’s participation in clinical research.

## 2. Methods

### 2.1 Procedures

The study used a survey methodology and was administered among a panel of adult survey takers in the US between January and March 2023. Survey respondents were recruited using Prolific (www.prolific.com), a survey provider that offers panel services to target specific groups in their database of respondents. For the current study, we selected respondents based on 3 criteria: sex at birth; age; and race, to obtain a sample that reflected the racial composition of the U.S. female population, as reported in the 2020 U.S. Census^31^. All participants provided informed consent prior to participating in the study and were compensated with $4 upon survey completion. The Mass General Brigham Institutional Review Board approved all procedures (IRB#2022P000169). Data are reported according to the American Association of Public Opinion Research (AAPOR) Transparency Initiative’s guidelines.

### 2.2 Participants

Women aged 18–70 years living in the U.S. and fluent in English were eligible to participate in the online survey.

### 2.3 Survey

The survey was administered in English and consisted of 81 questions divided into four sections: (1) demographics, (2) health status and current health experiences, (3) access to health-related information, and (4) attitudes and experiences related to clinical trials. Seventy-five questions were multiple-choice or a 5-point Likert scale, while 6 were open-ended (see *Supplementary Material*). Study data were collected and managed using REDCap electronic capture tools hosted by Mass General Brigham Research Applications team. REDCap (Research Electronic Data Capture) is a secure, web-based application designed to support data capture for research studies^32^. To ensure data quality, attention checks and human verification were embedded in the survey. The development of the survey began with a comprehensive literature review to identify previously validated questions that assessed relevant constructs, including health status, healthcare access, clinical trial awareness, and attitudes toward research participation. Content validity was assessed by three subject matter experts: a principal investigator with extensive clinical trial experience and two senior researchers with expertise in survey methodology. Additional questions were developed by the study team to assess participants’ awareness of sex-based differences related to health outcomes, testing of novel therapeutics and WTP. The validity of self-developed questions was tested via cognitive interviewing (see *Supplementary Material*).

### 2.4 Assessments

#### Sociodemographic characteristics

Demographic characteristics included age; sex at birth (female or male); race and ethnicity (race: Asian, Black or African American, White, or Other [Native Hawaiian or Other Pacific Islander, American Indian or Alaska Native]; ethnicity: Hispanic or non-Hispanic); educational attainment (high school graduate, college degree, graduate degree); annual household income; household composition (spouse/partner, children, other relatives); place of residence (urban, suburban, rural); US residency status; and primary language (English vs other).

#### Health status

To assess participants’ overall health, the survey used a validated global self-reported health status measure^33^. Participants were asked to rate their health on a 5-point scale ranging from “poor” to “excellent.” Additionally, nine other questions were included to evaluate participants’ past and current medical conditions, as well as their reproductive health history.

#### Healthcare access

An adapted version of the CIHI Patient Experiences Survey was used to assess participants’ access to healthcare services and types of barriers encountered^34^. Participants were asked how frequently, in the past 12 months, they were unable to access necessary healthcare services—such as lab tests or medications—due to financial barriers or life circumstances. Responses were recorded on a scale ranging from “never” to “very often.” A 3-point composite variable was created indicating whether respondents experienced poor, fair or good access to healthcare, based on (1) whether they had access to primary care and (2) whether they could use healthcare services needed.

#### Access to health-related information

An adapted version of the Survey of Health Information for Women was used to assess participants’ health information-seeking behavior^35^. This set of questions explored whether participants actively seek health-related information, their motivations for doing so, and the sources they rely on to obtain this information.

#### Interest in and attitudes toward clinical trial participation

Attitudes, knowledge and perceptions related to clinical trials were measured via questions extracted and adapted from three previously published assessment tools: the HealthStreet Health Needs Assessment (research perception)^36,37^, as modified by Otufowora and colleagues^25^, the Patient Attitudes toward Clinical Trial (PACT 22) scale^38^, and the Factors Influencing Participation in Clinical Trials^39^. Prior participation in clinical trials was assessed via questions developed by the Dana-Farber Cancer Institute Survey and Data Management Core to evaluate trial experiences in breast cancer patients^40^.

The HealthStreet Health Needs Assessment assesses the overall WTP in clinical trials, overall and by study type (minimal risk studies [e.g., surveys about participants’ health, review of medical records, giving a blood sample] and greater than minimal risk studies [having to take study medications, staying overnight in a hospital/clinic, giving a sample for genetic studies]). The PACT 22 scale consists of 22 statements that measure positive beliefs, safety, information needs, negative expectations, and patient involvement^38^. For each statement, respondents were asked if they agreed on a five-point Likert scale (1 = strongly agree to 5 = strongly disagree). Subscale scores were calculated by computing the means of each statement within the specific subscale. The Factors Influencing Participation in Clinical Trials scale assesses factors that represent a motivation to trial participation (0 = not motivating to 4 = most motivating), a barrier (0 = not a barrier to 4 = great barrier), or are considered helpful resources (0 = not helpful to 4 = most helpful)^39^.

#### Clinical trial experience

Prior participation in clinical trials was assessed via questions developed by the Dana-Farber Cancer Institute Survey and Data Management Core to evaluate trial experiences in breast cancer patients^40^. Whether they had been asked to participate in a CT in the past was measured and treated as a binary variable (Yes/No).

#### Awareness of female-specific factors

Sixteen questions were developed by the study team to assess participants’ awareness of sex-based differences, their views on the importance of examining such differences when evaluating the safety and efficacy of novel treatments, and the extent to which these considerations influence their willingness to participate (WTP) in clinical trials.

### 2.5 Statistical Analysis

Demographic, health, and clinical-trial–related variables were summarized overall and by race. Group differences in categorical variables were assessed with χ² tests: (1) White vs the combined racial-minority group and (2) among racial-minority subgroups (Black, Asian, and Other race). Racial differences in item-level clinical-trial variables (attitudes, motivating factors, barriers, decision-making) were examined with one-way analysis of variance (ANOVA).

The primary analysis modeled willingness to participate (WTP)—measured on a 3-point Likert scale (1 = not at all, 2 = maybe, 3 = definitely)—as a continuous outcome using multivariable linear regression. Predictors were race (White [reference], Black, Hispanic, Asian, Other race), age, educational attainment, annual household income, number of lifetime medical conditions, and access to health care (1 = low, 2 = moderate, 3 = high). We report unstandardized coefficients (β) with p values (and 95% CIs where available). The joint association of the race indicators was evaluated with a Type III (omnibus) F test (df = 3). As a secondary analysis, the same model was fit with self-reported health status as the outcome. Two-sided α = .05 defined statistical significance and all computations were performed in SAS, version 9.3 (SAS Institute Inc).

## 3. Results

### 3.1 Survey Sample

Table 1 summarizes participant characteristics overall and by race. A total of 5,301 respondents (99.5% cisgender women; mean [SD] age, 38 [13.6] years) completed the survey. Most were premenopausal (69.5%), had completed high school (40.3%), and lived with other adults (78.5%) and/or children younger than 18 years (80.4%). Approximately 30% reported annual household income <$40,000, and 51.4% resided in suburban areas. The majority spoke English as their first language (98%) and held U.S. citizenship (99.6%) (Table S1).

**Table 1.** Participant characteristics overall and by racial group.

| Characteristic | Overall<br>(n=5301)<sup>†</sup> | White<br>(n=3808) | Black<br>(n=715) | Asian<br>(n=364) | Other<br>(n=100) | +*p-value* | ++*p-value* |
| --- | --- | --- | --- | --- | --- | --- | --- |
| *Age (years)* |  |  |  |  |  | *0.0001* | *0.0001* |
| Mean (+/- SD) | 38.3 (13.6) | 39.6 (13.6) | 36.0 (13.2) | 29.8 (9.5) | 36.8 (13.0) |  |  |
| Range of Ages | 18-70 | 18-70 | 18-70 | 18-64 | 20-68 |  |  |
|  | **N (%)** | **N (%)** |  |  |  |  |  |
| *Gender* |  |  |  |  |  | *ns* | *ns* |
| Cisgender | 4926 (99.5) | 3766 (99.6) | 703 (99.4) | 358 (98.9) | 99 (99.0) |  |  |
| Transgender | 20 (0.4) | 14 (0.37) | 4 (0.57) | 1 (0.28) | 1 (1.0) |  |  |
| *Reproductive Age* |  |  |  |  |  | *0.0001* | *0.0001* |
| Pre-menopause | 3449 (69.5) | 2513 (66.3) | 534 (75.2) | 331 (91.4) | 71 (71.0) |  |  |
| Perimenopause | 450 (9.1) | 360 (9.5) | 65 (9.2) | 18 (5.0) | 7 (7.0) |  |  |
| Post-menopause | 1066 (21.5) | 920 (24.3) | 111 (15.6) | 13 (3.6) | 22 (22.0) |  |  |
| *Educational Attainment* |  |  |  |  |  | *0.026* | *0.0001* |
| Did not graduate high school | 33 (0.66) | 29 (0.77) | 3 (0.42) | 0 | 1 (1.0) |  |  |
| High school graduate and/or some college | 2004 (40.3) | 1500 (39.4) | 344 (48.3) | 102 (28.4) | 58 (58.0) |  |  |
| College graduate | 1922 (38.6) | 1471 (38.7) | 249 (35.0) | 172 (47.9) | 30 (30.0) |  |  |
| Additional coursework or graduate degree | 1015 (20.4) | 803 (21.1) | 116 (16.3) | 85 (23.7) | 11 (11.0) |  |  |
| *Annual Household Income* |  |  |  |  |  | *0.008* | *0.0001* |
| < \$40,000 | 1459 (30.3) | 1081 (29.1) | 264 (38.7) | 66 (20.4) | 48 (49.0) | | |
| \$40,000-\$70,000 | 1344 (27.9) | 1050 (28.2) | 192 (28.2) | 73 (22.5) | 29 (29.6) | | |
| \$70,001-100,000 | 984 (20.4) | 764 (20.5) | 136 (19.9) | 75 (23.2) | 9 (9.2) | | |
| > \$100,000 | 1036 (21.5) | 824 (22.2) | 90 (13.2) | 110 (33.9) | 12 (12.2) | | |
| *English is first language* | 4840 (98.0) | 3742 (99.3) | 705 (99.7) | 293 (80.5) | 100 (100) | *0.0001* | *0.0001* |

+*White* vs. *Non-White*, ++*Black* vs. *Asian* vs. *Other*
<sup>†</sup>*n=5301* represents the full sample, whereas *n=4987* had racial identity data available

Of the 5,301 respondents, 4,987 (94%) provided race information: 7% Asian, 14% Black, 2% Other race (Native Hawaiian or Other Pacific Islander, American Indian, or Alaska Native), and 77% White, mirroring the racial composition of U.S. women (2020 U.S. Census). White participants were older than participants from other racial groups (P < .001) and had higher educational attainment (P = .026) and greater annual household income (P = .008). They were also more likely to live alone (P = .008) and to reside outside urban areas (P = .0001).

Among racial minority groups, Asian women were younger (P < .001), had higher educational attainment, and were more likely to live in suburban areas (P < .001) compared with other racial minority participants. Respondents in the Other race group reported the lowest annual household incomes (49% earning <$40,000; P < .001).

### 3.2 Health Status

Most participants (69.6%) rated their health as good or very good, with no significant difference between White and racial minority participants (Table 2). More than 70% reported ≥2 lifetime medical conditions; the prevalence of ≥4 conditions were highest among White (36.6%) and Other race (47.0%) participants. Overall, the most frequently reported conditions were anxiety (61.7%), depression (57.5%), and migraine (38.3%). Brain-health conditions were the most reported category overall, followed by cardiovascular and gynecologic conditions. Among gynecologic conditions, fibroids (10.6%), endometriosis (8.0%), and infertility (6.5%) were most common; among cardiovascular conditions, hypertension (17.4%) and irregular heart rhythm (12.7%) predominated. Percentages reflect multi-response items.

**Table 2.**
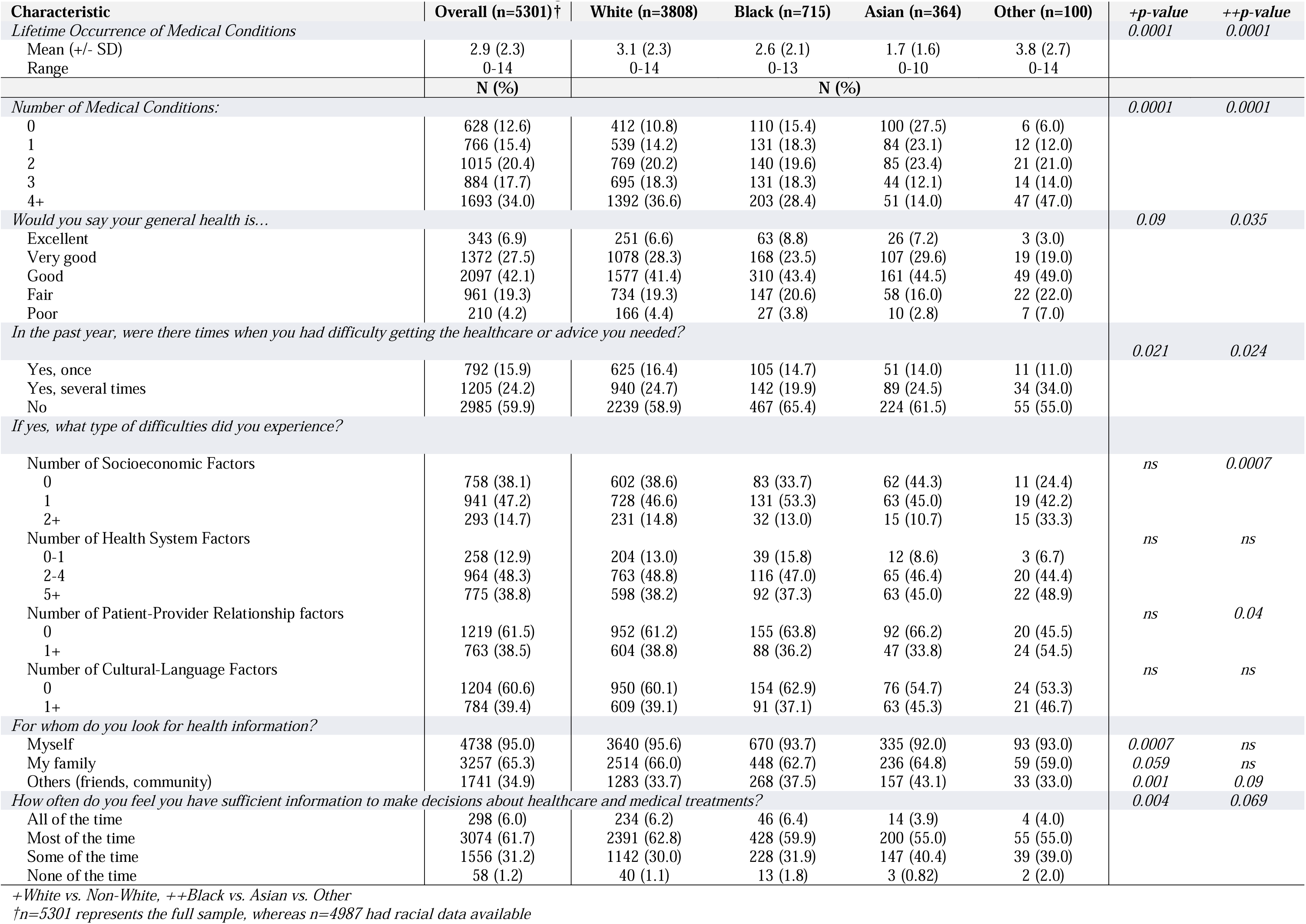
Health status, access to health care, and health information seeking behavior.

In multivariable analysis, older age (β = 0.003; 95% CI, 0.002–0.005; P = .0003), higher educational attainment (β = 0.059; 95% CI, 0.039–0.080; P < .0001), higher annual income (β = 0.033; 95% CI, 0.024–0.041; P < .0001), and better access to health care (β = 0.051; 95% CI, 0.018–0.085; P = .003) were associated with higher self-reported health, whereas a greater number of medical conditions was associated with lower health (β = −0.175; 95% CI, −0.185 to −0.164; P < .0001). Relative to White participants, Asian participants reported worse health (β = −0.190; 95% CI, −0.289 to −0.090; P = .002); estimates for Black (β = −0.033; 95% CI, −0.103 to 0.036) and Other race (β = −0.037; 95% CI, −0.205 to 0.131) were not significant (Table S3).

### 3.3 Access and Utilization of Health Care Services

Approximately 40% reported being unable to obtain needed health care at least once in the prior year. Among those reporting limited access, the highest proportion was in the Other race group (45%), followed by White (41%) and Asian (39%) participants. Differences in access were modest but statistically significant both for comparisons of racial minority groups with White participants and among minority groups (Ps < .05). The most common barriers were structural (appointment delays and geographic inaccessibility) (Table 2). Financial barriers (high out-of-pocket costs and lack of insurance coverage) were also frequently reported and were more prominent among Black and Other race participants than other groups (P = .0007).

### 3.4 Access to Health-Related Information

Most participants sought health information for themselves (95.0%) and for their families (65.3%) (Table 2). A majority reported having sufficient information to make health care decisions “most of the time” (61.7%) or “some of the time” (31.2%), with minor variability by race. Common sources were the internet (93.4%), physicians or other health professionals (88.2%), and family or friends (67.7%).

### 3.5 Attitudes Toward and Willingness to Participate (WTP) in Clinical Trials

Nearly 80% indicated they were “definitely” or “maybe” willing to participate in a clinical trial; 11% had ever been invited, and 7% had previously enrolled (Table 3). WTP was higher among White participants than among racial minority participants (P < .001). In multivariable linear regression, interest was lower among Black (β = −0.06; P = .04) and Asian (β = −0.09; P = .01) participants compared with White participants. Higher educational attainment (β = 0.03; P < .0001) and a greater number of medical conditions (β = 0.05; P < .0001) were associated with higher interest (Table 5). Within racial minority groups, Asian participants were least likely to report being “definitely” interested (12.9%) compared with Black (22.1%) and Other race (27.0%) participants (P = .0004).

**Table 3.**
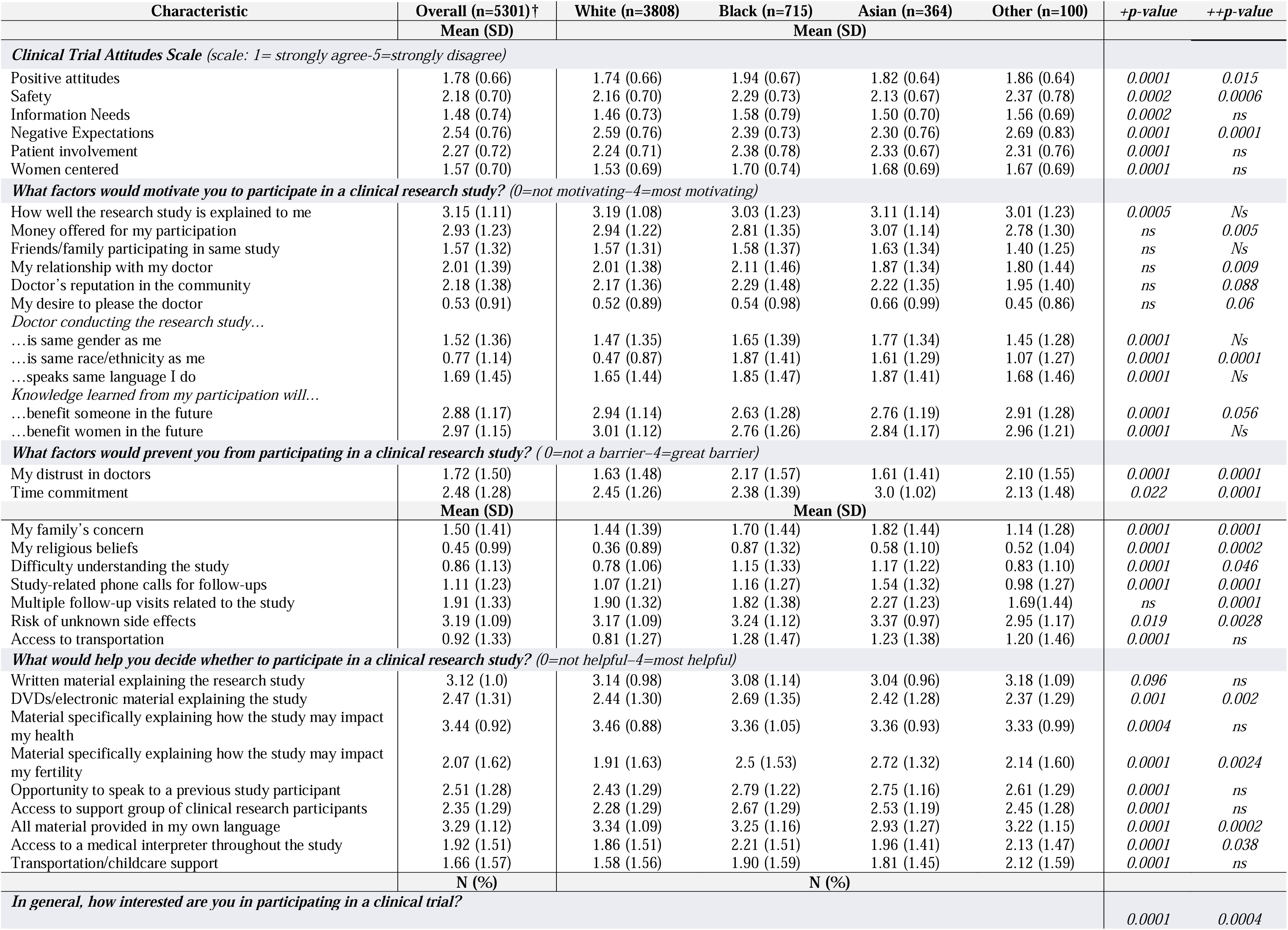
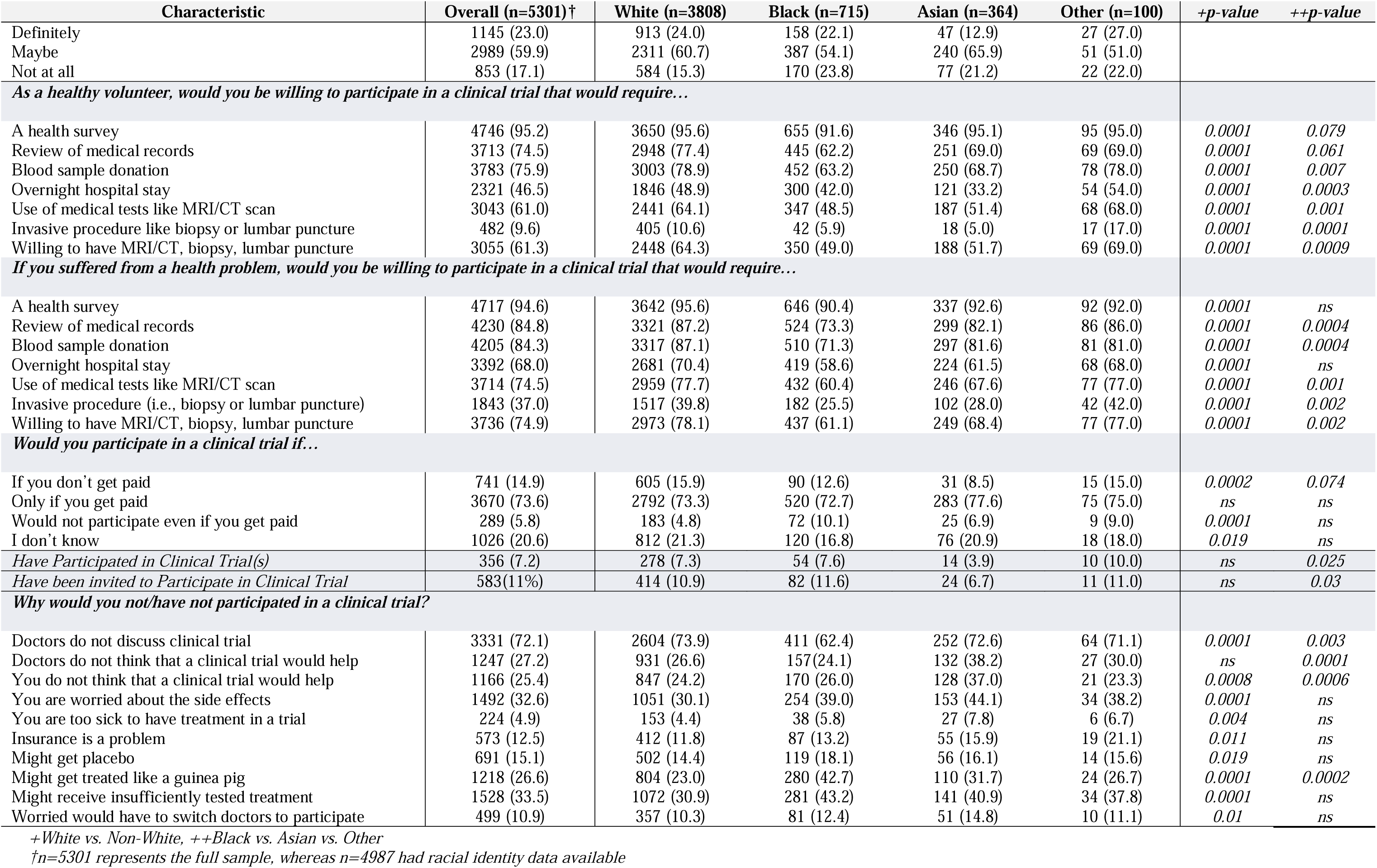
Clinical research perception and participation.

**Table 4.** Perceptions of sex differences.

| Characteristic | Overall (n=5301) †<br>N (%) | White (n=3769) | Black (n=702)<br>N (%) | Asian (n=363) | Other (n=100) | +p-value | ++p-value |
| --- | --- | --- | --- | --- | --- | --- | --- |
| *“It is important that women participate in clinical trials because their biology is different from men, thus they may respond differently to new treatments and/or medical techniques.”* |  |  |  |  |  | 0.0001 | ns |
| Strongly agree + agree | 4349 (87.6) | 3377 (89.2) | 585 (82.1) | 307 (84.8) | 80 (80.0) |  |  |
| Neutral | 467 (9.4) | 305 (8.1) | 101 (14.2) | 46 (12.7) | 15 (15.0) |  |  |
| Strongly disagree + disagree | 147 (3.0) | 106 (2.8) | 27 (3.8) | 27 (3.8) | 5 (5.0) |  |  |
| *How often do you feel that the health information you find applies specifically to you as a woman?* |  |  |  |  |  | 0.0001 | 0.0004 |
| 1: not at all | 72 (1.5) | 51 (1.3) | 16 (2.3) | 2 (0.55) | 3 (3.1) |  |  |
| 2-3: some of the time | 1293 (26.0) | 1031 (27.1) | 141 (19.9) | 99 (27.2) | 22 (22.5) |  |  |
| 4-6: most of the time | 3402 (68.4) | 2589 (68.1) | 492 (69.2) | 253 (69.5) | 69 (70.4) |  |  |
| 7: all of the time | 209 (4.2) | 133 (3.5) | 62 (8.7) | 10 (2.8) | 4 (4.1) |  |  |
| *How important is it for you that information about health, illnesses and treatments consider the specific biology and needs of women?* |  |  |  |  |  | ns | ns |
| 1: not at all | 49 (.99) | 38 (1.0) | 7 (1.0) | 4 (1.1) | 0 |  |  |
| 2-3: some of the time | 505 (10.2) | 384 (10.1) | 68 (9.6) | 46 (12.7) | 7 (7.1) |  |  |
| 4-6: most of the time | 2724 (54.8) | 2066 (54.4) | 397 (55.8) | 202 (55.8) | 59 (59.6) |  |  |
| 7: all of the time | 1693 (34.1) | 1311 (34.5) | 239 (33.6) | 110 (30.4) | 33 (33.3) |  |  |
| *For health problems (physical and/or mental/emotional) that affect men and women, do you think the symptoms and treatments are the same for both women and men?* |  |  |  |  |  | 0.0035 | ns |
| Yes to both | 510 (10.2) | 356 (9.4) | 93 (13.0) | 51 (14.0) | 10 (10.0) |  |  |
| Yes to one; ‘I don’t know’ to one | 800 (16.1) | 602 (15.8) | 130 (18.2) | 55 (15.11) | 13 (13.0) |  |  |
| No to one; ‘I don’t know’ to one | 1015 (20.4) | 785 (20.6) | 143 (20.0) | 66 (18.1) | 21 (21.0) |  |  |
| No to both | 1940 (38.9) | 1510 (39.7) | 248 (34.7) | 139 (38.2) | 43 (43.0) |  |  |
| I don’t know to both | 720 (14.4) | 553 (14.5) | 101 (14.1) | 53 (14.6) | 13 (13.0) |  |  |
| *How important is it to you that the safety and effectiveness of a medication or medical device is tested in both men and women?* |  |  |  |  |  | 0.025 | ns |
| It doesn’t matter | 301 (6.1) | 213 (5.6) | 58 (8.2) | 24 (6.6) | 6 (6.0) |  |  |
| It is preferable, but not necessary | 1915 (38.5) | 1489 (39.2) | 254 (35.7) | 134 (36.9) | 38 (38.0) |  |  |
| It is necessary | 2762 (55.5) | 2101 (55.3) | 400 (56.2) | 205 (56.5) | 56 (56.0) |  |  |
| *Would it be helpful to you if medication containers have a symbol indicating that it has been tested in women?* |  |  |  |  |  | 0.088 | 0.081 |
| Yes | 3428 (69.1) | 2595 (68.4) | 523 (73.6) | 237 (65.8) | 73 (73.0) |  |  |
| No | 800 (16.1) | 635 (16.7) | 96 (13.5) | 58 (16.1) | 11 (11.0) |  |  |
| I don’t know | 736 (14.8) | 563 (14.8) | 92 (12.9) | 65 (18.1) | 16 (16.0) |  |  |
| *Would it be helpful to you if instructions about use, dosage, or side effects of a medication contain information specific to women?* |  |  |  |  |  | ns | ns |
| Yes | 4400 (88.7) | 3352 (88.4) | 638 (89.7) | 319 (88.4) | 91 (91.9) |  |  |
| No | 283 (5.7) | 225 (5.9) | 39 (5.5) | 15 (4.2) | 4 (4.4) |  |  |
| I don’t know | 280 (5.6) | 215 (5.7) | 34 (4.9) | 27 (7.5) | 4 (4.0) |  |  |
| *When prescribed a medication or treatment, were you ever asked whether any of the following applied to you?* |  |  |  |  |  |  |  |
| Currently pregnant | 2364 (47.5) | 1882 (49.5) | 306 (42.9) | 125 (34.4) | 51 (51.0) | 0.0001 | 0.003 |
| Currently breastfeeding | 1618 (32.6) | 1304 (34.4) | 196 (27.5) | 82 (22.5) | 36 (36.0) | 0.0001 | 0.019 |
| Experiencing perimenopausal/menopause | 918 (18.4) | 732 (19.2) | 126 (17.7) | 38 (10.4) | 22 (22.0) | 0.008 | 0.0018 |
| Had a hysterectomy | 526 (10.6) | 425 (11.2) | 67 (9.4) | 18 (5.0) | 16 (16.2) | 0.012 | 0.0009 |
| Taking hormonal medications | 1155 (23.2) | 909 (23.9) | 134 (18.8) | 88 (24.4) | 24 (24.2) | 0.037 | 0.076 |

+White vs. Non-White, ++Black vs. Asian vs. Other
†n=5301 represents the full sample, whereas n=4987 had racial identity data available

**Table 5.** Associations of Race and Covariates with Willingness to Participate in Clinical Trials (Multivariable Linear Regression)

| Covariate | $\beta$ (95% CI) | *p*-value |
| --- | --- | --- |
| Age (years) | 0.000 (-0.001, 0.002) | *ns* |
| Educational Attainment | 0.034 (0.019, 0.049) | < 0.0001 |
| Household Income Range | -0.001 (-0.008, 0.005) | *ns* |
| Number of Medical Conditions | 0.054 (0.046, 0.062) | < 0.0001 |
| Access to Healthcare | -0.008 (-0.033, 0.017) | *ns* |
| Race (White=reference) |  |  |
| *Black* | -0.059 (-0.110, -0.008) | 0.024 |
| *Asian* | -0.090 (-0.163, -0.017) | 0.015 |
| *Other race\** | -0.067 (-0.191, 0.057) | *ns* |

\*Category includes Native Hawaiian or Other Pacific Islander, American Indian, or Alaska Native

Using the PACT-22 scale, participants generally endorsed positive attitudes toward clinical trials. Attitudes varied by race: White participants reported more positive perceptions than racial minority participants (P < .001), with Black participants reporting the lowest levels among minority groups. Most respondents preferred detailed information before enrollment (mean [SD], 1.48 [0.74]). While many agreed that safeguards protect participants (2.18 [0.70]), negative expectations were also common (2.54 [0.76]), particularly concerns about receiving placebo or less effective treatment. These concerns were more frequently endorsed by White participants than by racial minority participants (P < .0001), with the highest levels observed in the Other race group relative to other minority subgroups (P < .0001).

### 3.6 Factors Influencing Decision Making About Clinical Trial Participation

On the Factors Influencing Participation in Clinical Trials scale^39^, the most frequently endorsed motivators were helping others/benefiting other women (mean [SD], 2.97 [1.15]), receiving a clear explanation of the study (3.15 [1.11]), and financial compensation (2.93 [1.23]) (Table 3). White respondents rated clarity of study explanation and altruism as more influential than racial minority participants (differences modest; P < .0005). Among racial minority groups, Black women reported higher scores for factors related to the quality of the clinical team, including provider reputation and race-or language-concordance (P < .0001).

The most helpful resource for decision support was access to clear, language-concordant materials (mean [SD], 3.12 [1.00]), especially those addressing possible effects on health and fertility (3.44 [0.92]). Compared with White participants, racial minority women were more likely to identify peer education from prior participants as helpful.

The most frequently perceived barriers were potential study risks (i.e., side effects) and duration of participation. These concerns were more strongly endorsed by Asian respondents than by other racial groups (P = .0028). WTP was highest for minimal-risk studies (e.g., surveys; 95.2%) and lower for invasive procedures (9.6%) or extended time commitments (46.5%), although willingness increased when studies related to participants’ own health conditions. Overall, WTP was lower among racial minority women than White participants to participate either healthy volunteers or patients, with the exception of the Other race group, which reported greater willingness across study types (P = .0009).

### 3.7 Participation Experiences

Seven percent (n = 356) reported prior clinical trial participation: 4% Asian, 15% Black, 3% Other race, and 78% White (Table S2). More than 72% rated their experience as very good or excellent, with no significant racial differences. Positive aspects most often cited were financial compensation (28.6%), contributing to scientific advancement (22.4%), and access to health care services (14.6%). Negative aspects included time burden (23.3%), travel (15.4%), and procedure-related discomfort (13.2%). Sources of trial information were primarily online (65.7%) and health care providers (42.6%).

### 3.8 Women’s Inclusion in Clinical Trials and Consideration of Sex-Differences and Female-specific Factors

Nearly 88% of prior participants agreed that women’s participation in clinical trials is important because biological sex can influence treatment response (Table 4). Participants from racial minority groups were more likely than White participants to express neutrality on this statement (P < .0001). Consistent with these views, the majority of respondents strongly agreed that novel medications should be tested in both men and women (94%) and that drug labels should include sex-specific information (88.7%). Finally, Black and Asian women were less likely than White or Other race participants to report being asked about reproductive or hormonal status when prescribed a medication.

## Discussion

This study is among the first to systematically quantify how race, sociodemographic and economic factors, and health-related characteristics shape women’s engagement in clinical research. We surveyed a large sample stratified to mirror the racial composition of U.S. women, addressing external-validity concerns and enhancing generalizability. Our study shows that overall interest in clinical trials was high—nearly 80% of respondents were potentially interested—yet fewer than 11% had ever been invited and only 7% had enrolled, underscoring a substantial gap between interest and opportunity. Willingness to participate in clinical trials was mainly influenced by multimorbidity, followed by educational attainment and race. Notably, race also shaped the resources, motivators, and perceived barriers to participation, indicating the need for tailored engagement. Finally, awareness that sex differences affect health and treatment outcomes was high among our sample, suggesting that this knowledge is not confined to experts and may be harnessed to increase women’s engagement in clinical research.

In our cohort, education and multimorbidity were significantly associated not only with WTP in clinical trials but also with self-reported health, extending prior work that considered these outcomes separately. In particular, population-based surveys of U.S. adults have linked lower educational attainment to substantially worse self-reported health^41^, whereas longitudinal analyses of self-reported health trajectories in men and women indicate that, for women, improvements over recent decades are largely explained by gains in educational attainment^42^. In parallel, additional survey studies have associated higher education with greater WTP and better clinical-trial knowledge^43,44^. Several mechanisms may underlie these patterns: education is related to stronger health literacy, more effective navigation of health systems, greater access to supportive resources, and adoption of health-promoting behaviors—all of which can facilitate research engagement^45–47^. Indeed, participation in clinical trials has been shown to improve health outcomes, including all-cause and indication-specific mortality^48^. At the same time, education also shapes provider–patient interactions. Individuals with lower educational attainment—and, by extension, lower health literacy—may receive fewer trial invitations, reflecting both reduced access to services and clinician gatekeeping (e.g., assumptions about comprehension or protocol adherence)^49^. Such dynamics may disproportionately affect women and racial-minority groups^50–52^. Consistent with this, a small study reported that women— particularly those with limited health literacy—rely heavily on clinician recommendation when considering trial enrollment^53^.

When considering our sample, ∼41% of respondents had not graduated from college, ∼30% reported annual household income <$40,000, and ∼40% experienced difficulty obtaining needed care in the prior 12 months—reflecting combined financial and logistical barriers— despite most women reporting ≥2 lifetime medical conditions. Clear socioeconomic gradients by race were evident: Black and Other-race participants reported lower income and educational attainment than White women, whereas Asian women had the highest educational attainment and household income. Alongside education and multimorbidity, race had a statistically detectable but small association with self-reported health, reflecting its well-documented correlation with socioeconomic status and disease burden^54–56^. Asian participants reported lower health than White participants (β ≈ −0.19), suggesting that factors beyond SES (e.g., cultural norms in health reporting, health practices, and stress related to perceived or experienced discrimination in care) further influence how racial minorities evaluate their health^57,58^. Consistent with this interpretation, Asian respondents more often cited difficulties navigating the health system and language barriers as reasons for not obtaining needed care in the prior 12 months.

With regard to WTP, Asian and Black participants reported lower interest in trial participation than White participants, aligning with literature on mistrust, perceived risk, and structural exclusion from research among racial minorities^59,60^. Attitudes mirrored interest: White women held more positive views of trials, with Black women reporting the least favorable perceptions among minority groups. Willingness to participate was highest for low-risk studies and declined sharply with increasing invasiveness, though it increased again when participation was framed in the context of a personal health condition, consistent with our finding of a positive association between multimorbidity and WTP.

Beyond overall interest, our data offers a more nuanced understanding of the motivations and barriers influencing women’s decisions about clinical trial participation. Across racial groups, the most consistent facilitators were altruism (including benefiting other women), clear plain-language information, and appropriate compensation. Women from racial minority groups were especially likely to view peer education from prior participants and practical logistical supports (e.g., transportation, childcare) as helpful for decision-making. Notably, Black women placed greater weight on provider trustworthiness, reputation, and race- or language-concordance than other groups. This pattern aligns with prior evidence of lower trust in health-care providers among Black adults and points to the value of community-anchored approaches—such as peer ambassadors and partnerships with trusted organizations—to promote inclusive recruitment and retention^61^. Additionally, providing appropriate compensation mitigates financial obstacles to enrollment, which often fall most heavily on underrepresented populations such as women and racial/ethnic minorities^62^.

The most frequently cited barriers were the duration of participation—also among the top complaints from respondents with prior trial experience—and concerns about risk, reported most often by Asian women. Asian and Black respondents were also more likely to endorse difficulty understanding study materials and procedures. Taken together, these findings point to concrete levers for action: redesign study information for accessibility (plain-language, culturally and language-concordant), diversify dissemination channels (including peer education), and ensure content addresses issues that women prioritized—such as potential effects on fertility. These interventions should be coupled with burden-reducing supports—transportation/childcare assistance, flexible scheduling, and remote follow-up—which prior studies have also identified as important facilitators of women’s trial participation^16,20^. Furthermore, our results from the subgroup with trial experience underscore the importance of incorporating their voices into study design and operations.

Of note, this study adds insight into women’s views on their inclusion—and on the integration of sex-specific factors—in clinical research. Nearly nine in ten participants recognized women’s participation as critical given biological differences from men. Still, neutrality was more common among women from racially minoritized groups, indicating potential gaps in awareness or trust that may reflect prior exclusion or inadequate outreach. We also found a broad endorsement for sex-specific data, indicating that routine sex-disaggregated analyses and transparent sex-specific safety/efficacy reporting may increase trust and alignment with women’s needs. This finding suggest that the integration of sex in medical research and clinical practice is increasingly recognized among the lay public, and points to an additional pathway to enhance women’s engagement in research.

Our study is not without limitations. First, the cross-sectional design and reliance on self-reported measures limit causal inference and may introduce reporting bias, although several of our findings are consistent with prior reports. Second, only a small proportion of participants identified as Hispanic/Latina, which constrained our ability to examine the joint and independent contributions of race and ethnicity. Third, several racial identities were aggregated into an “Other race” category, reducing the granularity of race-specific estimates. Finally, we surveyed a broad population spanning many medical conditions rather than focusing on a single condition; this approach may obscure disparities specific to particular diseases, yet it provides a useful benchmark of U.S. women’s perceptions and attitudes toward clinical trials on which future, condition-specific studies can build.

Overall, U.S. women express strong interest in clinical research, but structural and informational barriers—disproportionately affecting racially minoritized women—constrain participation. Centering communication, trust, and burden reduction within community and clinical partnerships offers a practical path to translate interest into equitable enrollment and retention. At the system level, standardizing collection and discussion of reproductive/hormonal factors, and routinely disaggregating and reporting outcomes by sex, can advance equity and relevance of evidence for women.

## Acknowledgements

This work was supported by the Connors Center for Women’s Health Research Casey Toolin McAuliffe Memorial IGNITE Award, Brigham and Women’s Hospital, Boston, MA, USA. The research team would like to thank Anna Joseph for her help in data collection. We would also like to thank the Survey and Data Management Core at Dana-Farber Cancer Institute, and specifically Brett Nava-Coulter for his help in cognitive interviewing.

## Author Contributions Statement

PAS conceived the study, designed the study protocol and developed the survey with help from AR and RL. AR and JCW performed data collection. DF, TAR, AW and RL were responsible for data processing and data analysis. DF and PAS conceived the manuscript. All authors reviewed the manuscript and provided final approval of the version to be published.

## Data Availability

Data will be made available on request.

## References

1. Bernstein, S. R., Kelleher, C. & Khalil, R. A. Gender-based research underscores sex differences in biological processes, clinical disorders and pharmacological interventions. Biochem. Pharmacol. 215, 115737 (2023).

2. Bartz, D. et al. Clinical Advances in Sex- and Gender-Informed Medicine to Improve the Health of All: A Review. JAMA Intern. Med. 180, 574–583 (2020).

3. National Academies of Sciences, Engineering, and Medicine; Health and Medicine Division; Board on Population Health and Public Health Practice; Committee on Assessment of NIH Research on Women’s Health. Overview of Research Gaps for Selected Conditions in Women’s Health Research at the National Institutes of Health: Proceedings of a Workshop— in Brief. (National Academies Press (US), Washington (DC), 2024).

4. Regensteiner, J. G. et al. Barriers and solutions in women’s health research and clinical care: a call to action. Lancet Reg. Health Am. 44, 101037 (2025).

5. Studies, I. of M. (US) C. on E. and L. I. R. to the I. of W. in C., Mastroianni, A. C., Faden, R. & Federman, D. NIH Revitalization Act of 1993 Public Law 103-43. in Women and Health Research: Ethical and Legal Issues of Including Women in Clinical Studies: Volume I (National Academies Press (US), 1994).

6. National Institutes of Health. NIH Guidelines on the inclusion of women and minorities as subjects in clinical research. Fed. Regist. 59, 1408–1413 (1994).

7. Report of the Fourth World Conference on Women: Beijing, 4-15 September 1995. (United Nations, New York, 1996).

8. NOT-OD-15-102: Consideration of Sex as a Biological Variable in NIH-funded Research. https://grants.nih.gov/grants/guide/notice-files/not-od-15-102.html.

9. U.S. Food and Drug Administration. Guideline for the study and evaluation of gender differences in the clinical evaluation of drugs; notice. Fed. Regist. 58, 39406 (1993).

10. S. Department of Health and Human Services, Food and Drug Administration. Enhancing the Diversity of Clinical Trial Eligibility Criteria, Enrollment Practices, and Trial Design: Guidance for Industry. https://www.fda.gov/regulatory-information/search-fda-guidance-documents/enhancing-diversity-clinical-trial-eligibility-criteria-enrollment-practices-and-trial-design (2020).

11. Steinberg, J. R. et al. Analysis of Female Enrollment and Participant Sex by Burden of Disease in US Clinical Trials Between 2000 and 2020. *JAMA Netw*. Open 4, e2113749 (2021).

12. Sosinsky, A. Z. et al. Enrollment of female participants in United States drug and device phase 1-3 clinical trials between 2016 and 2019. Contemp. Clin. Trials 115, 106718 (2022).

13. Zhou, S., Qi, K., Nugent, B. M., Bersoff-Matcha, S. J. & Struble, K. Participation of HIV-1 infected treatment-naive females in clinical trials and sex differences in efficacy and safety outcomes. AIDS Lond. Engl. 37, 895–903 (2023).

14. Carcel, C. et al. Representation of Women in Stroke Clinical Trials: A Review of 281 Trials Involving More Than 500,000 Participants. Neurology 97, e1768–e1774 (2021).

15. Mayor, J. M. et al. Persistent under-representation of female patients in United States trials of common vascular diseases from 2008 to 2020. J. Vasc. Surg. 75, 30–36 (2022).

16. Bierer, B. E., Meloney, L. G., Ahmed, H. R. & White, S. A. Advancing the inclusion of underrepresented women in clinical research. Cell Rep. Med. 3, 100553 (2022).

17. Daitch, V. et al. Underrepresentation of women in randomized controlled trials: a systematic review and meta-analysis. Trials 23, 1038 (2022).

18. Avery, E. & Clark, J. Sex-related reporting in randomised controlled trials in medical journals. The Lancet 388, 2839–2840 (2016).

19. Sugimoto, C. R., Ahn, Y.-Y., Smith, E., Macaluso, B. & Larivière, V. Factors affecting sex-related reporting in medical research: a cross-disciplinary bibliometric analysis. The Lancet 393, 550–559 (2019).

20. Hawke, L. J. et al. Influences on clinical trial participation: Enhancing recruitment through a gender lens - A scoping review. Contemp. Clin. Trials Commun. 38, 101283 (2024).

21. Igwe, E. et al. Patient perceptions and willingness to participate in clinical trials. Gynecol. Oncol. 142, 520–524 (2016).

22. Jacobson, M. H. et al. Understanding willingness and barriers to participate in clinical trials during pregnancy and lactation: findings from a US study. BMC Pregnancy Childbirth 24, 504 (2024).

23. Ding, E. L., Powe, N. R., Manson, J. E., Sherber, N. S. & Braunstein, J. B. Sex Differences in Perceived Risks, Distrust, and Willingness to Participate in Clinical Trials: A Randomized Study of Cardiovascular Prevention Trials. Arch. Intern. Med. 167, 905–912 (2007).

24. Cardenas-Rojas, A. et al. Factors influencing clinical trial participation of women with fibromyalgia across the United States: a cross-sectional survey. Women Health 64, 369–379 (2024).

25. Otufowora, A. et al. Sex Differences in Willingness to Participate in Research Based on Study Risk Level Among a Community Sample of African Americans in North Central Florida. J. Immigr. Minor. Health 23, 19–25 (2021).

26. Lee, G. E., Ow, M., Lie, D. & Dent, R. Barriers and facilitators for clinical trial participation among diverse Asian patients with breast cancer: a qualitative study. BMC Womens Health 16, 43 (2016).

27. Tu, S.-P. et al. Clinical trials: Understanding and perceptions of female Chinese-American cancer patients. Cancer 104, 2999–3005 (2005).

28. Bontemps-Jones, J. E. et al. Beyond Tuskegee: A contemporary qualitative assessment of barriers to research participation among Black women. Cancer 131, e35648 (2025).

29. Markan, U. et al. Psychosocial Factors That Influence a Woman’s Decision to Enroll in a Clinical Trial: Implications on How to Improve Clinical Trial Enrollment Among Black Women. Int. J. Radiat. Oncol. Biol. Phys. 119, 1347–1356 (2024).

30. Khadraoui, W., Meade, C. E., Backes, F. J. & Felix, A. S. Racial and Ethnic Disparities in Clinical Trial Enrollment Among Women With Gynecologic Cancer. *JAMA Netw*. Open 6, e2346494 (2023).

31. Bureau, U. C. 2020 Census Demographic Profile. Census.gov https://www.census.gov/data/tables/2023/dec/2020-census-demographic-profile.html.

32. Harris, P. A. et al. Research electronic data capture (REDCap)—A metadata-driven methodology and workflow process for providing translational research informatics support. J. Biomed. Inform. 42, 377–381 (2009).

33. Strawbridge, W. J. & Wallhagen, M. I. Self-Rated Health and Mortality Over Three Decades: Results from a Time-Dependent Covariate Analysis. Res. Aging 21, 402–416 (1999).

34. Wong, S. T. & Haggerty, J. Measuring Patient Experiences in Primary Health Care: A Review and Classification of Items and Scales Used in Publicly-Available Questionnaries. (Centre for Health Services and Policy Research, Vancouver, BC, 2013).

35. Warner, D. & Procaccino, J. D. Toward wellness: Women seeking health information. J. Am. Soc. Inf. Sci. Technol. 55, 709–730 (2004).

36. Translational Medicine - What, Why and How: An International Perspective.

37. Cottler, L. B., Striley, C. W., O’Leary, C. C., Ruktanonchai, C. W. & Wilhelm, K. A. Engaging the Community in Research with the HealthStreet Model: National and International Perspectives. https://karger.com/books/book/237/chapter/5162998/Engaging-the-Community-in-Research-with-the.

38. Jenkinson, C. et al. Patient attitudes to clinical trials: development of a questionnaire and results from asthma and cancer patients. Health Expect. 8, 244–252 (2005).

39. Kurt, A. et al. Racial Differences Among Factors Associated with Participation in Clinical Research Trials. J. Racial Ethn. Health Disparities 4, 827–836 (2017).

40. Catalano, P. J. et al. Representativeness of participants in the cancer care outcomes research and surveillance consortium relative to the surveillance, epidemiology, and end results program. Med. Care 51, e9–15 (2013).

41. Borgonovi, F. & Pokropek, A. Education and Self-Reported Health: Evidence from 23 Countries on the Role of Years of Schooling, Cognitive Skills and Social Capital. PLoS ONE 11, e0149716 (2016).

42. Hill, T. D. & Needham, B. L. Gender-Specific Trends in Educational Attainment and Self-Rated Health, 1972–2002. Am. J. Public Health 96, 1288–1292 (2006).

43. Kim, J. Y. et al. The influence of socioeconomic status on individual attitudes and experience with clinical trials. Commun. Med. 4, 172 (2024).

44. Langford, A. T., Orellana, K. T. & Buderer, N. Correlates of knowledge of clinical trials among U.S. adults: Findings from the 2020 Health Information National Trends Survey. Contemp. Clin. Trials 114, 106676 (2022).

45. Viinikainen, J. et al. Does better education mitigate risky health behavior? A mendelian randomization study. Econ. Hum. Biol. 46, 101134 (2022).

46. van der Heide, I. et al. The Relationship Between Health, Education, and Health Literacy: Results From the Dutch Adult Literacy and Life Skills Survey. J. Health Commun. 18, 172– 184 (2013).

47. Cutler, D. M. & Lleras-Muney, A. Education and Health: Evaluating Theories and Evidence. https://www.nber.org/papers/w12352 (2006) doi:10.3386/w12352.

48. Baker, J. R. et al. Clinical trial participation improves outcome: A matched historical cohort study. Clin. Trials 10, 735–743 (2013).

49. Niranjan, S. J. et al. Bias and stereotyping among research and clinical professionals: Perspectives on minority recruitment for oncology clinical trials. Cancer 126, 1958–1968 (2020).

50. Grammoustianou, M. et al. Tackling healthcare providers bias: a systematic review of interventions with implications for inclusive clinical research. eClinicalMedicine 89, 103513 (2025).

51. Dorsey, B. F. et al. Health Care Provider Bias in Estimating the Health Literacy of Caregivers in a Pediatric Emergency Department. Pediatr. Emerg. Care 39, e80–e85 (2023).

52. Kim, I., Field, T. S., Wan, D., Humphries, K. & Sedlak, T. Sex and Gender Bias as a Mechanistic Determinant of Cardiovascular Disease Outcomes. Can. J. Cardiol. 38, 1865– 1880 (2022).

53. Burks, A. C., Doede, A., Showalter, S. L. & Keim-Malpass, J. Perceptions of Clinical Trial Participation Among Women of Varying Health Literacy Levels. Oncol. Nurs. Forum 47, 273–280 (2020).

54. Poverty Rate by Race/Ethnicity | KFF State Health Facts. KFF https://www.kff.org/state-health-policy-data/state-indicator/poverty-rate-by-raceethnicity/.

55. Quiñones, A. R. et al. Racial and Ethnic Differences in Multimorbidity Changes over Time. Med. Care 59, 402–409 (2021).

56. Williams, D. R., Mohammed, S. A., Leavell, J. & Collins, C. Race, Socioeconomic Status and Health: Complexities, Ongoing Challenges and Research Opportunities. Ann. N. Y. Acad. Sci. 1186, 69–101 (2010).

57. Population, C. on, Education, C. on B. and S. S. and, Education, D. of B. and S. S. and & Council, N. R. Racial and Ethnic Differences in the Health of Older Americans. (National Academies Press, 1997).

58. Kessler, R. C., Mickelson, K. D. & Williams, D. R. The Prevalence, Distribution, and Mental Health Correlates of Perceived Discrimination in the United States. J. Health Soc. Behav. 40, 208–230 (1999).

59. Broadening clinical trial participation to improve health equity. Deloitte Insights https://www.deloitte.com/us/en/insights/industry/health-care/increasing-diversity-clinical-trials.html.

60. George, S., Duran, N. & Norris, K. A Systematic Review of Barriers and Facilitators to Minority Research Participation Among African Americans, Latinos, Asian Americans, and Pacific Islanders. Am. J. Public Health 104, e16–e31 (2014).

61. LaVeist, T. A., Nickerson, K. J. & Bowie, J. V. Attitudes about Racism, Medical Mistrust, and Satisfaction with Care among African American and White Cardiac Patients. Med. Care Res. Rev. 57, 146–161 (2000).

62. Bierer, B. E., White, S. A., Gelinas, L. & Strauss, D. H. Fair payment and just benefits to enhance diversity in clinical research. J. Clin. Transl. Sci. 5, e159 (2021).

