## Supplementary Material for "Understanding Barriers to Clinical Trial Participation Among U.S. Women: A National Survey Study"

**By**

**Doruntina Fida, MPH^1,2^, Therese A. Rajasekera, PhD^1-3^, Ancella Roy, MSc^4^, Julia C. Wilson, BA^5^, Aleta Wiley, MPH^1,2^, Ruth Lederman, MPH^6,^ Primavera A. Spagnolo, MD PhD^1-3^**

1: Connors Center for Women’s Health Research, Brigham and Women’s Hospital, Boston, MA, USA

2: Department of Psychiatry, Brigham and Women’s Hospital, Boston, MA, USA

3: Harvard Medical School, Boston, MA, USA

4: University of Massachusetts Chan Medical School, Worcester, MA, USA

5: Columbia University Vagelos College of Physicians and Surgeons, New York, NY 10032

6: Survey and Data Management Core, Dana Farber Cancer Institute, Boston, MA

Correspondence should be addressed to:

Primavera A Spagnolo, MD, PhD

Mary Horrigan Connors Center for Women's Health Research

Dept. of Psychiatry, Brigham and Women’s Hospital

Harvard Medical School,

Thorn Building, 75 Francis Street

Boston, MA

**Supplementary Methods**

After drafting the survey instrument, the Survey and Data Management Core (SDMC) at Dana-Farber Cancer Institute was contracted to conduct a review on the design and usability of the survey instrument. After this review, the survey instrument was built into an online platform using REDCap. The SDMC then conducted cognitive interviews to ensure participant comprehension and uniform interpretation of the survey instrument. Since survey participants were recruited from a racially diverse array of US residents, cognitive interviews were performed with 11 women between the ages of 18-70 whose racial identities were representative of the survey population. The cognitive assessment protocol was reviewed and approved by the Mass General Brigham IRB. Upon obtaining informed consent, cognitive interviews were conducted remotely via video call or telephone by experienced interviewers at the SDMC and were designed to take no more than an hour to complete. Interviewers utilized a semi-structured interview style to elicit responses from participants. This consisted of first asking participants to complete the survey to assess understanding of survey questions. If any challenges in comprehension were identified, interviewers asked probing questions to clearly understand how survey questions were interpreted by the participants. Using this information, the survey instrument was revised, and participant follow-up was performed to ensure comprehension of the survey questions. Due to the length of the survey instrument, interviewers emphasized cognitive assessment on a limited number of specific questions. Of particular interest were questions regarding the inclusion of women in clinical trial participation and how women perceived their own health status.

**Supplementary Results**

| **Table S1.** Additional participant characteristics | | | | | | | |
| --- | --- | --- | --- | --- | --- | --- | --- |
| **Characteristic** | **Overall (n=5301)†** | **White (n=3808)** | **Black (n=715)** | **Asian (n=364)** | **Other (n=100)** | ***+p-value*** | ***++p-value*** |
|  | **N (%)** | **N (%)** | | | |  |  |
| *Household Composition* |  |  |  |  |  |  |  |
| *Living...* |  |  |  |  |  |  |  |
| Alone | 920 (21.5) | 680 (26.8) | 152 (26.8) | 69 (21.1) | 19 (23.2) | *0.008* | *ns* |
| With other adults | 3352 (78.5) | 2616 (79.4) | 415 (73.2) | 258 (78.9) | 63 (76.8) |  |  |
| With children ≤18 yrs. old | 1321 (80.4) | 1052 (80.6) | 181 (80.1) | 58 (81.7) | 30 (73.2) | *ns* | *ns* |
| With children >18 yrs. old | 323 (19.7) | 254 (19.5) | 45 (19.9) | 13 (18.3) | 11 (26.8) |  |  |
| *Residency Status* |  |  |  |  |  | *0.0001* | *ns* |
| US citizen | 4945 (99.6) | 3781 (99.8) | 707 (99.2) | 357 (98.1) | 100 (100) |  |  |
| *Location of residence* |  |  |  |  |  | *0.0001* | *0.0001* |
| Urban | 1555 (31.3) | 1052 (27.9) | 322 (45.3) | 152 (41.9) | 29 (29.0) |  |  |
| Suburban | 2551 (51.4) | 1973 (52.4) | 334 (50.0) | 197 (54.3) | 47 (47.0) |  |  |
| Rural | 833 (16.8) | 740 (19.7) | 55 (7.7) | 14 (3.9) | 24 (24.0) |  |  |
| *+White vs. Non-White, ++Black vs. Asian vs. Other* | | |  |  |  |  |  |
| *†n=5301 represents the full sample, whereas n=4987 had racial identity data available* | | | | | |  |  |

| **Table S2.** Additional sample characteristics and responses for those who participated in clinical trials (n=356) | | | | | | | |
| --- | --- | --- | --- | --- | --- | --- | --- |
| **Characteristic** | **Overall (n=356)** | **White (n=278)** | **Black (n=54)** | **Asian (n=14)** | **Other (n=10)** | ***+p-value*** | ***++p-value*** |
| ***Age (years)*** |  |  |  |  |  | *0.001* | *0.009* |
| Mean +/- SD | 43.9 (13.2) | 45.1 (13.4) | 42.1 (11.8) | 32.1 (6.1) | 36.7 (10.1) |  |  |
| Range of ages | 18-70 | 18-70 | 21-69 | 22-43 | 23-56 |  |  |
|  | **N (%)** | **N (%)** | | | |  |  |
| ***Educational Attainment*** |  |  |  |  |  | *0.019* | *0.013* |
| Did not graduate high school | 1 (0.28) | 0 | 0 | 0 | 1 (10.0) |  |  |
| High school graduate or some college | 103 (29.0) | 75 (27.1) | 25 (46.3) | 2 (14.3) | 1 (10.0) |  |  |
| College graduate | 136 (38.3) | 103 (37.2) | 21 (39.9) | 8 (57.1) | 4 (40.0) |  |  |
| Additional coursework/graduate degree | 115 (32.4) | 99 (35.7) | 8 (14.8) | 4 (28.6) | 4 (40.0) |  |  |
| ***Annual Household Income*** |  |  |  |  |  | *0.073* | *0.003* |
| < $40,000 | 105 (30.4) | 75 (27.7) | 24 (47.1) | 4 (28.6) | 2 (20.0) |  |  |
| $40,000-$70,000 | 81 (23.4) | 60 (22.1) | 13 (25.5) | 2 (14.3) | 6 (60.0) |  |  |
| $70,001-100,000 | 63 (18.2) | 53 (19.6) | 7 (13.7) | 2 (14.3) | 1 (10.0) |  |  |
| $100,000-$149,999 | 65 (18.8) | 57 (21.0) | 6 (11.8) | 1 (7.1) | 1 (10.0) |  |  |
| > $150,000 | 32 (9.3) | 26 (9.6) | 1 (2.0) | 5 (35.7) | 0 |  |  |
| ***Overall experience in a clinical trial*** | | | | | | *ns* | *ns* |
| Excellent | 131 (37.2) | 102 (37.2) | 17 (31.5) | 8 (57.1) | 4 (40.0) |  |  |
| Very good | 125 (35.5) | 99 (36.1) | 19 (35.2) | 3 (21.4) | 4 (40.0) |  |  |
| Good | 65 (18.5) | 46 (16.8) | 14 (25.9) | 3 (21.4) | 2 (20.0) |  |  |
| Fair | 19 (5.4) | 17 (6.2) | 2 (3.7) | 0 | 0 |  |  |
| Poor | 12 (3.4) | 10 (3.7) | 2 (3.7) | 0 | 0 |  |  |
| ***Which statement best describes the role you played when the decision was made about participating in a clinical trial?*** | | | | | | *ns* | *ns* |
| Made decision with little or no input from doctors | 235 (66.4) | 183 (66.1) | 35 (66.0) | 11 (78.6) | 6 (60.0) |  |  |
| Made decision after considering doctors' opinions | 68 (19.2) | 52 (18.7) | 11 (20.8) | 1 (7.1) | 4 (40.0) |  |  |
| Made decision together with doctor | 38 (10.7) | 31 (11.2) | 5 (9.4) | 2 (14.3) | 0 |  |  |
| Doctors made decision after considering your opinion | 7 (2.0) | 5 (1.8) | 2 (3.8) | 0 | 0 |  |  |
| Doctors made decision with little or no input from you | 6 (1.7) | 6 (2.2) | 0 | 0 | 0 |  |  |
| ***How did you learn that participating in a clinical trial was a possibility?*** | | | | | |  |  |
| Doctor/healthcare professional | 150 (42.6) | 119 (43.2) | 24 (45.3) | 4 (28.6) | 3 (33.3) | *ns* | *ns* |
| Family/friends | 85 (23.9) | 60 (21.6) | 17 (31.5) | 5 (35.7) | 3 (30.0) | *0.06* | *ns* |
| Media (TV, newspaper, internet) | 234 (65.7) | 178 (64.0) | 38 (70.4) | 10(71.4) | 8 (80.0) | *ns* | *ns* |
| Other | 60 (16.9) | 44 (15.8) | 10 (18.5) | 3 (21.4) | 3 (30.0) | *ns* | *ns* |
| ***What did you like the least about your participation?*** | | | | | |  |  |
| Large time commitment | 83 (23.3) | 65 (23.4) | 11 (20.4) | 5 (35.7) | 2 (20.0) | *ns* | *ns* |
| Travel/transportation issues | 55 (15.4) | 45 (16.2) | 8 (14.8) | 1 (7.1) | 1 (10.0) | *ns* | *ns* |
| Clinical activities | 47 (13.2) | 38 (13.7) | 6 (11.1) | 1 (7.1) | 1 (10.0) | *0.03* | *ns* |
| Other | 44 (12.3) | 36 (13.0) | 8 (14.8) | 0 | 0 | *ns* | *ns* |
| ***What did you like the most about your participation?*** | | | | | |  |  |
| Contributing to science/altruism | 80 (22.4) | 62 (22.3) | 10 (18.5) | 5 (35.7) | 2 (20.0) | *ns* | *ns* |
| Compensation | 102 (28.6) | 80 (29.5) | 17 (31.5) | 3 (21.4) | 0 | *ns* | *ns* |
| Receiving medical support/trial med. working | 52 (14.6) | 47 (16.9) | 5 (9.3) | 0 | 0 | *0.062* | *0.091* |
| Visits were informative/organized | 59 (16.5) | 41 (14.8) | 11 (20.4) | 1 (7.1) | 6 (60.0) | *ns* | *0.001* |
| *+White vs. Non-White, ++Black vs. Asian vs. Other* | | | | | | | |

**Table S3**. Associations With Self-reported Health Status (Multivariable Linear Regression)

| **Covariates** | **β (95% CI)** | ***p- value*** |
| --- | --- | --- |
| Age (years) | 0.003 (0.002, 0.005) | *0.0003* |
| Educational Attainment | 0.059 (0.039, 0.080) | *< 0.0001* |
| Household Income Range | 0.033 (0.024, 0.041) | *< 0.0001* |
| No. of Medical Conditions | -0.175 (-0.185, -0.164) | *< 0.0001* |
| Access to Healthcare | 0.051 (0.018, 0.085) | *0.003* |
| Race (White=reference) |  |  |
| *Black* | -0.033 (-0.103, 0.036) | *ns* |
| *Asian* | -0.190 (-0.289, -0.090) | *0.002* |
| *Other race** | -0.037 (-0.205, 0.131) | *ns* |
| **Category includes Native Hawaiian or Other Pacific Islander, American Indian, or Alaska Native* | | |

**Appendix A – Survey Script**

**Section 1. Current Health**

1. In general, compared to other people your age, how would you rate your overall health?

1. Excellent

2. Very Good

3. Good

4. Fair

5. Poor

2. Have you ever had any of the following physical and/or mental health conditions or related treatments? Mark: No, Yes, currently Yes in the past on each line.

Heart attack

Stroke

Hypertension

Heart failure

Irregular heart rhythm

Diabetes

Autoimmune diseases (rheumatoid arthritis, lupus, Crohn's disease, psoriasis)

Kidney failure or dialysis

Lung disease (not including cancer)

Osteoporosis

Migraine

Other neurological disorders

Cancer

Thyroid conditions

Endometriosis

Fibroids

Infertility

Liver disease/cirrhosis

Anxiety disorders (panic attack, generalized anxiety)

PTSD

Depression

Unipolar/bipolar disorder

- Psychosis disorder

3. Are you taking medications for any of these conditions? Yes/No /# meds:_
4. How would you describe your current menstrual status?
   - Premenopause (before menopause; having regular periods)

SKIP TO Q8

- Perimenopause/menopause transition (changes in periods, but have nt gone 12 months in a row without a period)
- Postmenopause (after menopause)

How was your menopause:

- Spontaneous (‘normal’)
- Surgical (removal of both ovaries)
- Due to chemotherapy/radiotherapy
- Other (explain):

5. Has your uterus been surgically removed?

Yes

No

I do not know

6. Have your ovaries been surgically removed?

No

Yes, both ovaries

Yes, one ovary or

don't know

.

7. If you are not having periods, how old were you when you last had regular period? (Your best guess.)

44 or younger, 45-49, 50-54, 55 or older

8. How many pregnancies lasting 6 months or more have you had?

0 1 2 3 4 5 6 7 8 or more

All referenced their own histories and felt certain of answer.

9. Which of the following are you currently using or have used previously (select all that apply):

Using now	Previously used

Birth control pill, ring, skin patch

IUD

Injectable hormone

Other

10. Have you ever used hormone therapy for menopause?

No

Yes, currently

Yes, in the past only

**Section 2. Seeking Health Information**

11. Where do you place yourself on a scale of those who seek information related to health (physical and/or mental/emotional)?7-point Likert scale; 1= I mostly try to avoid this type of information and 7= Very interested in this information
12. Here are some reasons people seek for information regarding health (physical and/or mental/emotional). Please indicate how often any of these reasons has applied/applies to you

☐ To understand how lifestyle habits, such as diet, physical exercise, smoking impact on my health

never, rarely, some of time, most of the time

☐ To understand if I have a health problem

never, rarely, some of time, most of the time

☐ To understand a health problem better

never, rarely, some of time, most of the time

☐ To feel more relaxed about a health problem

never, rarely, some of time, most of the time

☐ To find treatment options and plan courses of action for a health problem

never, rarely, some of time, most of the time

☐ I have never looked for health information	Skip to question 11

13. Here are some of the ways that people find information about health and health problems (physical and/or mental/emotional). Please check all you have used.

☐Family or friends

☐Doctors or others in the medical profession

☐Internet

☐ Books

☐ Magazines

☐ TV

☐Other: __________________

14. Overall, how useful are the health information you find from these sources?

7-point Likert scale; 1= Not at all useful and 7= Very useful

15. How easy or difficult is it to find health information?

☐Very easy

☐Somewhat easy

☐Somewhat difficult

☐Very difficult

☐Don’t know

16. For whom do you look for information (check all that apply)

- Myself
- Somebody else in my family
- Friends/acquaintances
- Community members

17. Does the information you find ever affect any of your decisions about health treatments or the way you care for someone else?

7-pointLikert scale; 1= not at all and 7= extremely

18. Have you ever changed health care decisions based on the information you found from the above sources? Yes/No
19. Does the information you get ever affect any of your decision about health treatments or the way you help care for someone else?

7-pointLikert scale; 1= not at all and 7= extremely

20. Did you ever seek health information because: [select all that apply]?

☐A medical professional recommended specific sources

☐You wanted to supplement the information given by a medical professional

☐You were not satisfied with the information given to you by a medical professional

☐There was not enough time to ask a medical professional more questions

☐You were hesitant to ask a medical professional any more questions

☐You wanted to find out more information on your own

☐You needed to make a decision about medical treatment

☐You wanted information about specific symptoms

☐You wanted information about disease prevention

☐You wanted information on alternative care

☐You wanted information for someone else

21. When looking for information about an illness, do you think that each of the following are addressed in a clear and through way : Yes, No, NA

- background of the condition
- symptoms
- treatment options

22. When looking for information about a medication or a drug, do you think that these points are clearly described? Yes, No, NA

- what the drug is used for
- dosage
- side effects
- possible interactions with other drugs you are taking
- whether the drug can be used while pregnant or breastfeeding

23. How often do you feel you have sufficient information to make decisions about healthcare and medical treatments?

- All of the time
- Most of the time
- Some of the time
- None of the time

24. On a scale from “not at all” to “very much”, how often do you feel that the health information you find applies specifically to you as a woman?

7-point Likert scale; 1= not at all and 7= very much

25. On a scale from “not at all” to “very much”, how important is it for you that information about health, illnesses and treatments consider the specific biology and needs of women?

7-point Likert scale; 1= not at all and 7= very much

26. Think about a medication that you have taken or are currently taking. How much does it matter to you that the medication was tested in both women and men before being available on the market?

7-point Likert scale; 1= not at all and 7= very much

27. For health problems (physical and/or mental/emotional) that affect both men and women, do you think that it is necessary that a medication or treatment is tested in both women and men?

- Yes
- No
- I don’t know

28. For health problems (physical and/or mental/emotional) that affect both men and women, do you think the SYMPTOMS are the same for both women and men?

- Yes
- No
- I don’t know

29. For health problems (physical and/or mental/emotional) that affect both men and women, do you think that TREATMENTS for those illnesses are the same for both women and men?

	Yes

	No

	I don’t know

30. Would you be willing to take a medication or use a medical device that has never been tested in women or was only tested in a small number of women?

	Yes

	No

	I don’t know

31. How important is it to you that the safety and effectiveness of a medication or a medical device is tested in both men and women?

- the gender does not matter as long as it gets tested
- it is preferable, but not necessary
- it is necessary

32. Would you be willing to use a medication that is less effective in women than men, but has fewer side effects?

	Yes

	No

	I don’t know

33. Would it be helpful to you if medication containers have a symbol indicating that it has been tested in women?

	Yes

	No

	I don’t know

34. Would it be helpful to you if the instructions about use, dosage, or side effects of a medication contain information specific to women?

	Yes

	No

	I don’t know

35. Has your doctor ever discussed benefits or side effects of a medication that are specific to women?

	Yes

	No

	I don’t know

36. Would you find it beneficial if your doctor gives you information about the dose and side effects of a medication that are specific to women?

	Yes

	No

	I don’t know

37. Please tell us in 1 or 2 sentences what health problems you would like to find more information on and why*(you can think about a health problem, a treatment, or a health condition such as pregnancy)*: ______________________________________________________
38. Please tell us in 1 or 2 sentences, for women in general, what is the most pressing health need you would like to see addressed and why: _______________________________________________________

### **Section 3. Current health experiences and behavior**

39. Do you have a regular clinic, doctor practice, or primary care doctor where you receive care?

Yes

No

40. How long does it take you to get to this provider’s office? ____ Mins and distance in Miles (approximate is fine)
41. How do you usually travel to this office (mode of transportation)? ________________

-personal vehicle (yours or someone else’s)

-public transportation (bus, train)

-hired vehicle (taxi, uber, lyft)

Other: _____________

42. In the past year, Were there times when you had difficulty getting the healthcare or advice you needed?

No -> Go to question 21 / Yes, once -> / Yes, several times

43. If yes, what type of difficulties did you experience?

Difficulty contacting a physician / A specialist was unavailable / Difficulty getting an appointment / Do not have personal/family physician / Waited too long to get an appointment / / Service not available at time required / Service not available in my area / Transportation problems / Cost issues / Language barriers / Did not feel comfortable with the available doctor or nurse / Did not know where to go / Unable to leave the house because of a health problem / Other, please specify: _____

44. “Was there a time in the past 12 months when you needed to see a doctor but could not because of cost?” Yes/No
45. In the past 12 months, Were there times when you did not take medicines prescribed by a doctor because of their costs? Never / Rarely / Sometimes / Often / Very often / I don’t take any drugs
46. In the past 12 months, were there times when you did not get laboratory tests or exams because of their costs? Never / Rarely / Sometimes / Often / Very often / I did not get laboratory tests or exams
47. In the past 12 months, were there times when you did not get services recommended by your doctor that aren’t covered by health insurance because of their costs? (such as physiotherapy, psychotherapy, nutrition counseling…) Never / Rarely / Sometimes / Often / Very often / I did not use these services
48. In the past 12 months, were there times when you found it difficult to get health care because you had to take time off work? Never / Rarely / Sometimes / Often/ Very often
49. In the past 12 months, were there times when you found it difficult to get health care services because of the additional costs it involves? (babysitting, parking, etc.) Never / Rarely / Sometimes / Often/ Very often / I did not need these services
50. When prescribed a medication or treatment were you ever asked whether any of the following applied to you (for each answer yes or no)?

Currently Pregnant

Currently Breastfeeding

Experiencing Perimenopausal/menopause

Had a Hysterectomy

Taking hormonal medications

51. In general, how much do you pay attention to how the products you buy affect your health?

7-point Likert scale; 1= totally disagree and 7= totally agree

52. Have you ever stopped taking a medication because it was difficult to take it at the frequency or time prescribed?

Yes

No

**Section 4. Clinical Research Perception and Participation**

53. In general, how interested are you in participating in a clinical trial?

☐Definitely/Maybe ☐Not at all

54. What factors would motivate you to participate in a clinical research study? *Please rate the following from Not Motivation (0) to MOST (4) motivating*

My relationship with my doctor 0,1,2,3,4

Doctor’s reputation in the community 0,1,2,3,4

How well the research study is explained to me 0,1,2,3,4

My desire to please the doctor 0,1,2,3,4

Money offered for my participation 0,1,2,3,4

A friend or family member participating in the same study 0,1,2,3,4

The doctor conducting the research is the same gender (sex) as of me 0,1,2,3,4

The doctor conducting the research is the same race/ethnicity as me 0,1,2,3,4

The doctor conducting the research speaks the same language as I do 0,1,2,3,4

Knowledge learned from my participation will benefit someone in the future 0,1,2,3,4

Knowledge learned from my participation will benefit other women in the future 0,1,2,3,4

55. What factors would prevent you from participating in a clinical research study? Please rate the following from NOT A BARRIER (0) to GREAT (4) barrier.

My distrust in doctors

Time commitment

My family’s concern

My religious beliefs

Clinical research studies are too hard to understand

Study related phone calls for follow-ups

Multiple follow-up visits related to the study

Risk of unknown side effects

Access to transportation

Other: _________________________

56. What would help you decide whether to participate in a clinical research study? *Please rate the following from NOT HELPFUL (0) to MOST HELPFUL(4) helpful resource*

Written material explaining the research study

DVDs or electronic material explaining the research study

Having material specifically explaining how the study may impact my fertility

Having the opportunity to speak to a patient who has participated in a clinical research study

Having access to a support group of patients who have participated in clinical research

Having all material provided in my own language

Having access to a medical interpreter throughout the study

Having transportation/childcare support

Other: __________________________

*Clinical Trial Attitudes Scale*

57. Please tell us whether you agree or disagree with the following statements.

- a. New and better treatments can only be produced if patients agree to take part in clinical trials
- b. It is important that women are adequately included in clinical trials
- c. Without the results from clinical trials, doctors would be less able to select the best treatment
- d. Pharmaceutical companies should ensure that valid clinical trials are conducted on every drug before it is generally available
- e. If most patients refused to take part in clinical trials, important developments in medicine would be seriously delayed.
- F. Clinical trials are carried out according to strict rules to safeguard the interests of patients.
- G. I assume that drug treatments that have been prescribed for me have already been thoroughly tested in clinical trials in other women.
- H. Clinical trials are only conducted on drugs for which there is already evidence to show that they are likely to be effective.
- i. The conduct of all clinical trials is carefully regulated to ensure that the results are valid.
- J. I would want as much written information as possible about a clinical trial before I agreed to take part in a clinical trial.
- K. I would want to know if I would be likely to get side effects by taking part in a clinical trial before I agreed to take part.
- L. I would only take part in a clinical trial if I thought I understood everything about it.
- M. I think I would find being in a clinical trial frightening.
- N. I would only take part in the clinical trial if I thought that my own health would benefit.
- O. I would only take part in a clinical trial if I thought that I would not be inconvenienced by doing so.
- P. I would only take part in a clinical trial if I knew which treatment I am going to receive.
- Q. I would only take part in a clinical trial if I was sure that the doctor treating me knew which treatment I was getting.
- R. If I was satisfied with my current drug treatment, I would probably refuse to take a different drug in a clinical trial.
- S. It is important for people to take part in clinical trials to confirm the value of new treatments and / or medical techniques.
- T. I would take part in a clinical trial because the results should benefit patients like me in the future.
- U. I think all patients who are eligible should be asked to take part in clinical trials.
- V. It is important that women participate in clinical trials because their biology is different from men, thus they may respond differently to new treatments and / or medical techniques.
- W. Unless advised by their doctor to withdraw from a trial, all patients should cooperate fully

58. Have you ever participated in a clinical trial?

Yes

No

If you answer Yes, please respond the questions 7-11. If you answered No, please go to question 12

59. In general, how would you rate your overall experience?

- Excellent
- Very Good
- Good
- Fair
- Poor

60. Please describe what you liked the MOST about your clinical trial participation:___________
61. Please describe what you liked LEAST about your clinical trial participation:______________
62. How did you learn that participating in a clinical trial was a possibility? Please mark yes or no for each item.

☐From a doctor or other healthcare professional?

☐ From a family member?

☐From a friend or acquaintance?

☐From a patient support or advocacy group?

☐Read in newspaper, magazine, or other publication?

☐Heard on radio or saw on television?

☐Internet (website, chat room, discussion board)?

☐From some other source (please specify):___________________

63. Which statement best describes the role you played when the decision was made about participating in a clinical trial?

☐ You made the decision with little or no input from your doctors

☐ You made the decision after considering your doctors’ opinions

☐ You and your doctors made the decision together

☐ Your doctors made the decision after considering your opinion

☐ Your doctors made the decision with little or no input from you

64. Why didn’t you participate in a clinical trial? Please mark yes or no for each item.

☐a. Your doctors did not discuss clinical trial options with you.

☐b. Your doctors did not think that a clinical trial would help you.

☐c. You did not think that a clinical trial would help you.

☐d. You were worried about the side effects of the treatment in the clinical trial.

☐e. You were too sick to have treatment in a clinical trial.

☐f. Insurance coverage or payment was a problem.

☐g. You might get placebo or sugar pill rather than actual treatment.

☐h. You might get treated like a “guinea pig”.

☐i. You might receive treatment that had not been sufficiently tested.

☐j. You were worried that you would have to switch doctors in order to participate in a clinical trial.

☐k. Other reason (please specify):_________________________

65. As a healthy volunteer, would you be willing to participate in a clinical trial that requires any of these? Please check all that apply

☐ a health survey

☐Review of medical records

☐blood sample donation

☐ an overnight hospital stay

☐use of medical tests like MRI/CT scan

☐invasive procedure like biopsy or lumbar puncture

66. If you were to think about participating in a clinical trial for treating a disease or health problem that you suffer from, would you be willing to participate in a clinical trial that requires any of these? Click all that apply

☐ a health survey

☐Review of medical records

☐blood sample donation

☐ an overnight hospital stay

☐use of medical tests like MRI/CT scan

☐invasive procedure like biopsy or lumbar puncture

67. Would you participate in a clinical trial if?

- if you don’t get paid
- only if you get paid
- Would Not participate even if you get paid
- I don’t Know

68. Before completing this survey, is there anything that you would like to comment on or add relating to your health, health needs and/or healthcare use as a woman?

___________________________________________________________________________

69. Is there anything you would like to comment about or add about clinical research participation?

_____________________________________________________________________________

### **Demographics:**

70. What is your sex as assigned at birth?

Female

Male

Prefer not to answer

71. What is your age?

☐______ (in Years)

☐ Not sure

☐ Refuse to Answer

72. What is the highest grade you finished in school?

☐ Less than 9 years of school

☐ Some high school (9-11 years)

☐ High school graduate, or GED

☐ Some college or technical school

☐ College degree graduate

☐ Graduate degree

☐ Post graduate education, but no higher degree

☐ I prefer not to answer

73. **Which of these is the highest level of education you have completed?**

☐ No formal qualifications

☐ Secondary education (e.g. GED/GCSE)

☐ Secondary education (e.g. GED/GCSE)

☐ Technical/community college

☐ Undergraduate degree (BA/BSc/other)

☐ Graduate degree (MA/MSc/MPhil/other)

☐ Doctorate degree (PhD/other)

☐ Don't know / not applicable

74. With whom do you live? *(Mark all that apply with an X.)*

☐ Spouse / partner

☐ Parents/ parents-in-law

☐ Girlfriend / boyfriend

☐ Live alone

☐ Children aged 18 years or younger

☐ Children over aged 18 years

☐ Other relative, specify: ________

☐ Others, specify: _____________

75. What is your current annual household income?

☐ Less than $40,000

☐ $40,000-49,999

☐ $50,000-$59,000

☐ $60,000-$69,000

☐ $70,001-$79,999

☐ $80,001-$89,999

☐ $90,001-$99,999

☐ $100,000- $149,999

☐ More than $150,000

☐ I prefer not to answer

76. Do you live in the United States of America: Yes, No
77. Is English your first language?

☐ Yes

☐ No

7A. If no, what is your first language? _______________________________

78. Do you consider yourself to be Hispanic or Latina? Hispanic, Spanish, or Latina is a person of Mexican, Puerto Rican, Cuban, South or Central American, or other Spanish culture of origin, regardless of race.

☐ Yes, Hispanic or Latina

☐ No, not Hispanic or Latina

79. Which of the following would you use to describe yourself? *Check all that apply.*

☐ Native Hawaiian or other Pacific Islander

☐ American Indian or Alaska Native

☐ Asian

☐ Black or African American

☐ White

☐ Other (write in) _____________

80. Which of the following describes your residency status?

"U.S. citizen,"

"permanent resident,"

"work permit,"

"student visa,"

"other."

81. Which one of the following best describes the location of where you live?

☐ Urban (you live in a city)

☐ Suburban (you live in a suburb of a city, within 30 miles of a city)

☐ Rural (you live outside the city, in the countryside, over 30 miles from a city)

☐Other (please describe) ______
